# Use of illness severity scores to predict mortality in interstitial lung disease patients hospitalised with acute respiratory deterioration

**DOI:** 10.1101/2022.07.11.22277492

**Authors:** Rachel L Williams, Catherine Hyams, Joe Robertshaw, Maria Garcia Gonzalez, Zsuzsa Szasz-Benczur, Paul White, Nick A Maskell, Adam Finn, Shaney L Barratt, the AvonCAP Research Group

**Affiliations:** Academic Respiratory Unit, University of Bristol, North Bristol NHS Trust, Southmead, Bristol, BS10 5NB; Research and Innovation, North Bristol NHS Trust, Southmead, Bristol BS10 5NB; Bristol Interstitial Lung Disease Service, North Bristol NHS Trust, Southmead, Bristol BS10 5NB; Bristol Vaccine Centre, Schools of Population Health Sciences and Cellular and Molecular Medicine, University of Bristol, Bristol, BS2 8AE; Vaccine and Testing Team, UHBW NHS Trust, Bristol; University of the West of England, Bristol, BS16 1QY

**Author notes:** These authors contributed equally to this work (Rachel L Williams, Catherine Hyams). **Correspondence to:** Dr Shaney Barratt, Bristol Interstitial Lung Disease Service, North Bristol NHS Trust, Southmead Hospital, Bristol BS10 5NB, UK.

**Keywords:** Interstitial lung disease, acute respiratory deterioration, severity scores, pulmonary fibrosis

## Abstract

**Introduction:** Hospitalisations relating to acute respiratory deteriorations (ARD) in Interstitial Lung Disease (ILD) have poor outcomes. Factors predicting adverse outcomes are not fully understood and data addressing the use of illness severity scores in prognostication are limited.

**Objective:** To investigate the use of CURB-65 and NEWS-2 severity scores in the prediction of mortality following ARD-ILD hospitalisation, using prospective methodology and to validate previously determined cut-offs, derived from a retrospective study cohort.

**Methods:** A dual-centre prospective observational cohort study of all adults (≥18y) hospitalised with ARD-ILD in Bristol, UK (n=179). Gender-Age-Physiology (GAP), CURB-65 and NEWS-2 scores were calculated for each eligible admission.

Receiver operating characteristics (ROC) curve analysis was used to quantify the strength of discrimination for NEWS-2 and CURB-65 scores. Univariable and multivariable logistic regression analyses were performed to explore the relationship between baseline severity scores and mortality.

**Results:** GAP showed some merit at predicting 30-day mortality (AUC=0.64, *P*=0.015); whereas CURB-65 showed modest predictive value for in-hospital (AUC=0.72, *P*<0.001) and 90-day mortality (AUC=0.67, *P*<0.001). NEWS-2 showed higher predictive value for in-hospital (AUC=0.80, *P*<0.001) and 90-day mortality (AUC=0.75, *P*<0.001), with an optimal derived cut-off ≥6.5 found to be sensitive and specific for predicting in-hospital (83% and 63%) and 90-day (73% and 72%) mortality. In exploratory analyses, GAP score addition improved the predictive ability of NEWS-2 against 30-day mortality and CURB-65 across all time-periods.

**Conclusion:** NEWS-2 has good discriminatory value for predicting in-hospital mortality and moderate discriminatory value for predicting 90-day mortality. The optimal NEWS-2 cut-off value determined was the same as in a previous retrospective cohort, confirming the NEWS-2 score shows promise in predicting mortality following ARD-ILD hospitalisation.

## INTRODUCTION

Idiopathic pulmonary fibrosis (IPF) is considered the archetypal chronic progressive fibrotic Interstitial Lung Disease (ILD), which can have an unpredictable clinical course punctuated by sudden, severe acute respiratory deteriorations (ARD). ARD related hospitalisations in IPF patients are associated with poor patient outcomes, posing a significant burden on healthcare services.^1–4^ ARD-IPF are categorised into parenchymal (e.g. pneumonia) and extra-parenchymal causes (e.g. pleural effusion) within a conceptual framework.^1^ Acute exacerbation (AE), defined within this framework describes rapid respiratory deterioration associated with new widespread parenchymal ground glass opacification (with or without consolidation) on the background of established fibrosis and is not fully explained by fluid overload or cardiac failure. Mortality associated with AE-IPF is high,^1,5^ although, factors predicting adverse outcomes are not fully understood. Whilst originally described in IPF, both ARD and AE are increasingly considered to be features of other fibrosing ILDs; although, ARD and AE epidemiology and triggers in non-IPF ILDs are less well understood, with no randomised controlled trials examining optimal management.^5^

The emergence of SARS-CoV-2 and resultant pandemic have severely impacted healthcare provision, with ILD patients at increased risk of severe COVID-19 disease, possibly attributable to immunosuppressive treatment in addition to their chronic lung changes.^6–8^ Ensuring appropriate healthcare resource allocation and usage, especially with the significant ILD disease burden, may be aided by routinely used and validated illness scores. The CURB-65 score is a national standard and was validated against 30-day mortality in pneumonia,^9^ and subsequently validated against other conditions including sepsis. The National Early Warning Score-2 (NEWS-2) is used routinely throughout UK hospitals to rapidly identify patients at risk of deterioration,^10^ and also to predict in-hospital mortality. Both these scores are undertaken routinely with no requirement for invasive or additional testing and are thus widely available to physicians. There are, however, few data on the use of admission illness severity scores to predict short to medium term outcomes following admission with ARD.^11–13^

Our previous single-centre, retrospective, observational cohort study of 172 IPF patients admitted with ARD, supported the use of CURB-65 and NEWS-2 illness severity scores to predict in-hospital and 90-day mortality.^4^ This analysis identified baseline CURB-65□>□3.5 and NEWS-2 score >6.5 as the optimum cut-offs for predicting in-hospital mortality. ARD-IPF mortality was high irrespective of cause, in-line with mortality estimates from other published cohorts;^3^ however, data also suggested AE-IPF mortality was higher compared to other parenchymal causes of ARD.^14^

The AvonCAP prospective observational study of acute lower respiratory tract disease (aLRTD) provided the opportunity to describe the characteristics of a prospective cohort of patients hospitalised with ARD-ILD following SARS-CoV-2 emergence. We therefore aimed to pragmatically evaluate whether admission CURB-65 and NEWS-2 scores could be used as predictors of mortality following ARD-ILD requiring hospitalisation, as these practical scores which are routinely used may aid both specialists and non-specialists in optimising treatment for patients with ILD. We then compared patients following an episode of ARD requiring hospitalisation with IPF and non-IPF ILDs and investigated factors associated with worse outcome.

## MATERIALS AND METHODS

### Study Design

A prospective, dual-centre observational cohort study undertaken at North Bristol and University Hospitals Bristol and Weston NHS Foundation Trusts, encompassing all secondary care institutions in Bristol, UK, as part of the AvonCAP study. The study was approved by the Health Research Authority East of England Ethics Committee, including Section 251 of the 2006 NHS Act approved by the Confidentiality Advisory Group (REC:20/EE/0157, ISRCTN:17354061).

### Study Subjects

All admission lists were screened by members of the clinical care team to identify patients hospitalised with worsening respiratory signs/symptoms between 1^st^ August 2020 and 9^th^ November 2021. Full inclusion and exclusion criteria are available on IRSCTN (Supplementary Data 1).^15^ Only individuals with a multidisciplinary team (MDT)-confirmed diagnosis of ILD (either pre-existing or arising from the hospitalisation), including but not limited to IPF, were included in this analysis.

Collection of clinical data was undertaken on all eligible participants using a standardised REDCap proforma^16^. Lung Function Tests (LFTs) and 6-min walk test (6MWT) were included if conducted within 6-months of hospitalisation and were not performed specifically for this study. Gender-Age-Physiology (GAP), CURB-65 and NEWS-2 scores at the point of admission and Charlson Comorbidity Index (CCI), which is a predictor of 10-year mortality, were calculated for each hospital admission (Supplementary Data 2).^9,17–19^ Mortality data (30- and 90-day mortality) was collected through linked national healthcare data records using a unique identifier given to each patient.

### Case Definitions

The aetiology of ARD-ILD was categorised according to Collard’s conceptual framework:

(1) Extra-parenchymal causes: pleural effusion, pneumothorax or pulmonary embolism,

(2) AE-ILD: diagnosed in accordance with broadened Collard *et al*. revised criteria ^1^ to include all fibrosing ILDs: previous/concurrent ILD, worsening dyspnoea <1 month duration, new bilateral ground-glass opacification (with/without consolidation) on CT imaging, superimposed on background of established fibrosis and not fully explained by cardiac failure/ fluid overload. AE-ILD was further sub-categorised into triggered (clear precipitant) or idiopathic.

(3) Not AE-ILD: other parenchymal ARD-ILD causes not attributed to an AE, including: non-pneumonic lower respiratory tract infection (NP-LRTI)/presumed NP-LRTI (defined in the context of a CT not clearly identifying a radiological cause for the deterioration or unchanged CXR (presumed) but CRP>6mg/ml); pneumonia; cardiac failure/fluid overload; disease progression; and, those with non-specific trigger (no radiological cause demonstrated on CT imaging with CRP<6mg/ml; for example: anxiety, symptom control and/or palliation). Respiratory infection, regardless of pathogen, was categorised into pneumonic or NP-LRTI – thus COVID-19 was classified as a ‘Not AE-ILD’ cause of ARD.

(4) Not fully classified ARD-ILD: hospitalisation without CT imaging on admission but unaltered chest radiograph and CRP <6mg/ml and no clear trigger.

### Outcome Measures

The primary outcome of this analysis was to validate previously determined admission (i.e. at the time of hospitalisation) CURB-65 score ≥3.5 and NEWS-2 score ≥6.5 as predictors of in-hospital mortality^4^ in a broader group of patients with ILD.

Secondary outcomes were to determine the utility of the GAP score to predict in-hospital, 30- and 90-day mortality rates in patients with ARD-ILD. We also determined the overall in-hospital, 30- and 90-day mortality rates for patients with ARD-ILD, hospital length of stay (LOS), thereby highlighting any differences in mortality between IPF and non-IPF ARD cohorts.

### Statistical Analysis

Categorical data were presented as numbers and proportions (n, %), continuous non-parametric data as medians and interquartile range (IQR). Either log rank test, Fisher’s exact test or chi-square test was used where appropriate to analyse differences between groups.

For the primary analysis, univariable and multivariable logistic regression analyses were performed to explore the relationship between baseline severity scores and mortality. The factors used in the multivariable model were decided *a priori* and were smoking status, GAP score, CURB-65 and NEWS-2 score. Receiver operating characteristic (ROC) curve analysis was used to quantify the strength of discrimination. Previous data suggested cut-offs of CURB-65>3.5 and NEWS-2>6.5, provided Area Under the Receiver Operating Curve (AUROC) estimates of 0.85 and 0.89 respectively, in the prediction of in-hospital mortality in hospitalised ARD-IPF.^4^ Assuming an inpatient mortality rate approximating 20%.^4^ a consecutive sample size of 175 patients would be sufficient to validate an AUROC≥0.8 with a lower 95% confidence interval (CI) of the AUROC exceeding a lower acceptability threshold of 0.7.^20^ Power calculations were performed using the proprietary Power Analysis & Sample Size (PASS) software. The impact of prognostic factors on survival was computed using a Kaplan-Meier analysis and multivariate Cox proportional hazards model. For all tests, a *P*<0.05 was considered statistically significant. Data were analysed using IBM SPSS v28.0.

## RESULTS

### Patient Demographics

Of the total 122,097 patients ≥18 years hospitalised in the study period: 179 patients had confirmed ARD with a multidisciplinary diagnosis of ILD (Figure 1A). The median age of patients was 75 years (IQR 72-84), 64% were male and 57% were ex-smokers (Table 1). IPF was the underlying diagnosis of 39% of the cohort, but a broad range of other ILD diagnoses existed, including: unclassifiable ILD (13%), hypersensitivity pneumonitis (HP) (12%), and connective tissue disease associated-ILD (10%) (Table 1). *De novo* presentations with ILD were infrequent (3% ILD admissions). Patients had moderately restrictive disease (median FVC% predicted 75 (IQR 63-91), TLCO 44% (IQR 33-58) and a median GAP score of 4 (IQR 3-5), corresponding to GAP stage II. Approximately one third of patients had at least 2 or more (31%, n=56) concurrent medical co-morbidities. Overall, vaccination rates were high in the cohort: 75% of patients having received pneumococcal and seasonal influenza vaccination by the time of admission, and 83% of eligible patients (Supplementary Data 3) having received at least one dose of a COVID-19 vaccination prior to admission. The median length of stay was 7 days (IQR3-11) versus 6 days (IQR 3-11) for IPF and non-IPF ARD admissions (*P*=0.545), respectively.

**Figure 1.**
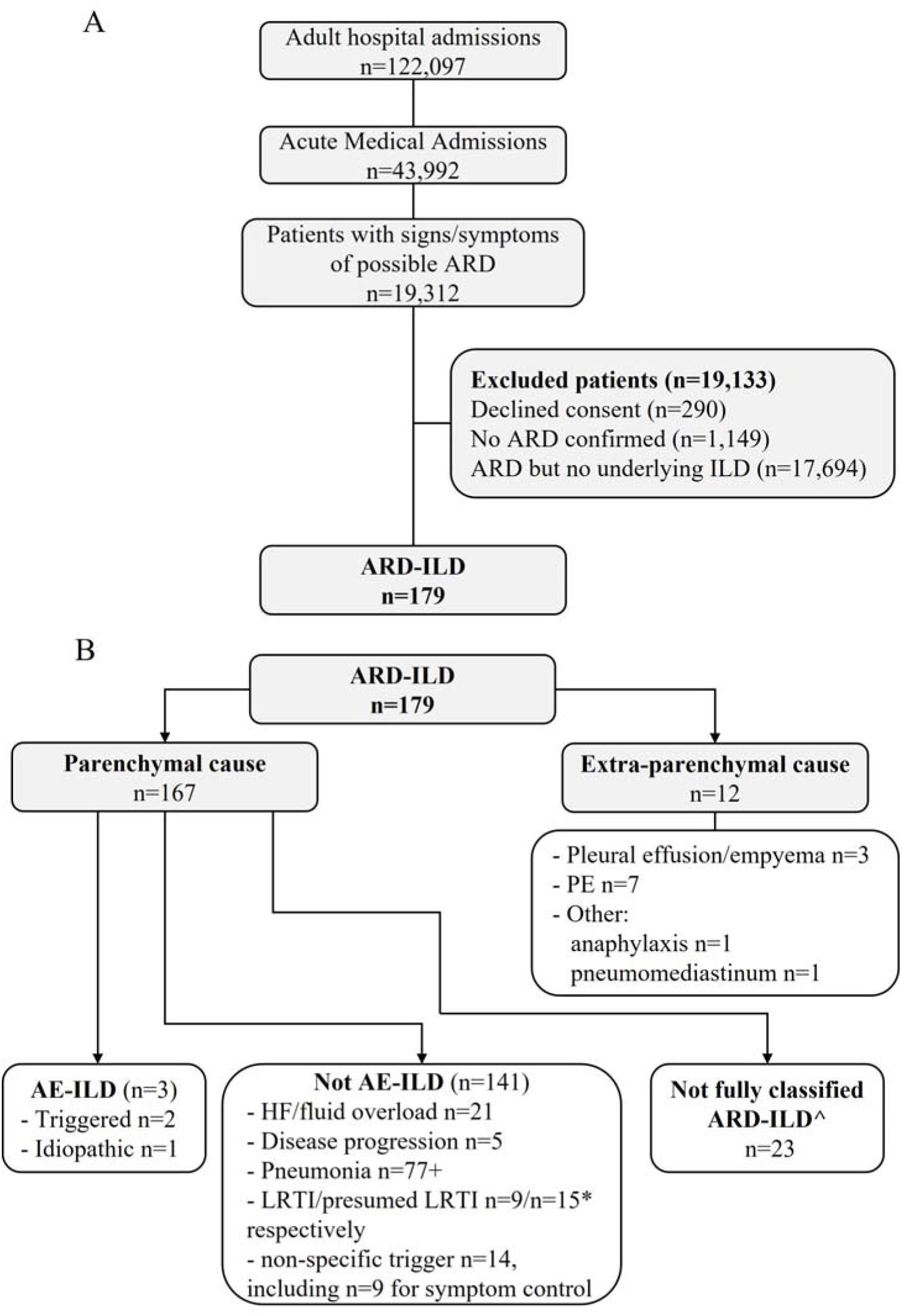
Adults hospitalised with acute respiratory deterioration of interstitial lung disease. (A) Flow diagram of study participants and (B) aetiology of acute respiratory deterioration of Interstitial lung disease (ARD-ILD). ARD; acute respiratory deterioration; AE-ILD, acute exacerbation of interstitial lung disease; LRTI, lower respiratory tract infection; COVID-19, coronavirus disease 2019; HF, heart failure. +n=11 COVID-19 pneumonia, one aspiration pneumonia, one hospitalised pneumonia, remainder deemed to be community acquired pneumonia *including 3 with symptomatic COVID-19 but no CXR infiltrates ^ unclassified – CT not performed and CRP <50.

**Table 1:** Characteristics of adults hospitalised with ARD-ILD

| Characteristic | ARD-ILD<br>n = 179 | IPF<br>n = 70 | Non IPF<br>n = 109 | P-value |
| --- | --- | --- | --- | --- |
| **Male, n (%)** | 114 (64) | 54 (77) | 60 (55) | **0.0040** |
| **Age (years), median (IQR)** | 79 (72-84) | 81 (75-86) | 77 (70-83) | **0.0068** |
| **Smoking Status, n (%)** |  |  |  | 0.2537 |
| Current | 9 (5) | 4 (6) | 5 (5) |  |
| Ex-smoker | 102 (57) | 48 (69) | 54 (50) |  |
| Never | 53 (30) | 14 (20) | 39 (36) |  |
| Unknown | 15 (8) | 4 (6) | 11 (10) |  |
| **Ethnicity, n (%)** |  |  |  | 0.7566 |
| Caucasian | 148 (83) | 58 (83) | 90 (83) |  |
| Other | 11 (6) | 6 (9) | 5 (5) |  |
| Unknown | 20 (11) | 6 (9) | 14 (13) |  |
| **CCI, median (IQR)** | 5 (4-6) | 5 (4-6) | 5 (3-6) | 0.8765 |
| **Aetiology ILD, n (%)** |  |  |  | N/A |
| IPF | 70 (39) | 70 (100) | - |  |
| Unclassifiable | 23 (13) | - | 23 (13) |  |
| Hypersensitivity pneumonitis | 22 (12) | - | 22 (12) |  |
| CTD-ILD | 17 (10) | - | 17 (10) |  |
| Smoking related ILD | 14 (8) | - | 14 (8) |  |
| CPFE (IPF with emphysema) | 9 (5) | - | 9 (5) |  |
| NSIP | 8 (5) | - | 8 (5) |  |
| Sarcoid | 6 (3) | - | 6 (3) |  |
| Asbestosis | 4 (2) | - | 4 (2) |  |
| Other □ | 6 (3) | - | 6 (3) |  |
| **Baseline PFTs, median (IQR)** |  |  |  |  |
| **Lung function** |  |  |  |  |
| FEV1 % predicted | 78 (66-93), n=168 | 81 (69-93), n=68 | 75 (63-93), n=100 | 0.5627 |
| FVC % predicted | 75 (63-91), n=168 | 75 (64-89), n=68 | 77 (63-96), n=100 | 0.5324 |
| TLCO % predicted | 44 (33-58), n=137 | 40 (30-46), n=58 | 54 (37-61), n=79 | **0.0002** |
| 6MWT distance (m) | 240 (155-311), n=133 | 240 (130-290), n=60 | 245 (168-354), n=73 | 0.7552 |
| 6MWT minimum sats (%) | 87 (84-90) n=133 | 87 (84-89), n=60 | 87 (84-90), n=73 | 0.8129 |
| **Total GAP score** | 4 (3-5), n=150 | 5 (4-6), n=61 | 4 (3-5), n=89 | **<0.0001** |
| **Admission Severity Scores** |  |  |  |  |
| CURB-65, median (IQR) | 2 (1-4) | 2 (1-4) | 1 (1-3) | **0.0096** |
| NEWS-2, median (IQR) | 6 (3-8) | 6 (3-8) | 5 (2-8) | 0.4246 |
| **ARD-ILD cause** |  |  |  | 0.8284 |
| Extra-Parenchymal | 12 (7) | 5 (7) | 7 (6) |  |
| AE-ILD | 3 (2) | 2 (3) | 1 (1) |  |
| Not AE-ILD | 141 (79) | 53 (76) | 88 (81) |  |
| Pneumonia | 77 (43) | 33 (47) | 44 (40) |  |
| Not fully classified ARD-ILD | 23 (13) | 9 (13) | 14 (13) |  |
| **Respiratory Support, n (%)** |  |  |  |  |
| None | 20 (11) | 10 (14) | 10 (9) | 0.3346 |
| Oxygen | 159 (89) | 60 (86) | 99 (91) |  |
| HFNO | 20 (11) | 8 (11) | 12 (11) | 1.0000 |
| NIV | 8 (4) | 5 (7) | 3 (3) | 0.2123 |
| IMV | 1 (1) | 0 (0) | 1 (1) | 0.4033 |
| **Treatment, n (%) #** |  |  |  |  |
| **Antifibrotic on admission** | 40 (22) | 40 (57) | 0 (0) | N/A |
| Received antibiotics (>2 days) | 139 (78) | 53 (74) | 86 (79) | 0.4391 |
| Received corticosteroids | 49 (27) | 18 (26) | 31 (28) | 0.7702 |
| **Length of stay, median (IQR)** |  |  |  |  |
| Hospitalisation days | 6 (3-11) | 7 (3-11) | 6 (3-11) | 0.5451 |
| **Mortality, n (%)** |  |  |  |  |
| In-hospital mortality | 34 (19) | 17 (24) | 17 (16) | 0.1848 |
| 30-day mortality | 42 (23) | 21 (30) | 21 (19) | 0.0901 |
| 90-day mortality | 69 (39) | 33 (47) | 36 (33) | 0.0614 |
| Survival days, median (IQR) | 59 (10-144), n=101 | 23 (10-23), n=41 | 79 (13-144), n=60 | **0.0417** |

*P*-values represent the result of a Log-rank Test or Fisher's exact test between IPF and non-IPF groups
□ Other includes lymphocytic interstitial pneumonia, Langerhan's cell histiocytosis, pleuro-parenchymal fibroelastosis, Rosai-Dorfman, organising pneumonia.

IPF patients were statistically older than those with non-IPF ILD diagnoses (IPF 81 years vs non-IPF 77 years, *P*=0.0068), more likely to be male (IPF 77% vs non-IPF 55%, Fisher’s exact test, *P* =0.004), with lower TLCO % predicted (IPF 38% vs non-IPF 54%, *P*=0.0002) and higher GAP scores (IPF 5 vs non-IPF 4, *P*<0.0001) on hospitalisation. IPF and non-IPF ILD patients had comparable baseline spirometry values (Table 1).

### Underlying Aetiology

Most (79%) ARD-ILD admissions were due to parenchymal causes other than AE-ILD (Figure 1B). Pneumonia was the most common parenchymal cause (57%, 77/141); the vast majority were community-acquired (83% of pneumonias, n=63), with COVID-19 pneumonitis in eleven patients (14%). Other parenchymal causes included cardiac failure (n=21, 15%), NP-LRTI (n=24, 17%, including 3 patients with symptomatic COVID) and disease progression in n=5 (4%). An extra-parenchymal pathology was considered the cause of ARD in 12 patients (7%); 7 with pulmonary embolism, 3 with pleural effusion/empyema, with pneumomediastinum and anaphylaxis in the remaining patients.

Non-specific triggers of ARD were identified in approximately 10% of patients and these admissions were related to requirements for palliation and symptom control, including anxiety and breathlessness. AE-ILD was rare in this cohort (n=3, 2%) and seen in patients with fibrotic hypersensitivity pneumonitis and IPF. Due to low incidence of AE-ILD we were not able to perform analyses to determine differences between patients admitted with AE-ILD and other parenchymal causes.

### Primary Outcome

Over half of all patients hospitalised with an ARD-ILD had a CURB-65≥2 (n=99, 55%) and NEWS-2 ≥5 (n=114, 64%) on admission. Patients with ARD-IPF had a statistically higher baseline CURB-65 compared to those with non-IPF ARD-ILD diagnoses (median CURB-65 2 [IQR 1-4] versus 1 [IQR 1-3], *P*=0.0096), but baseline NEWS-2 scores were comparable (median NEWS-2 6 [IQR 3-8] versus 5 [IQR 2-8] respectively, *P* >0.05) (Table 1).

GAP score was found to have diminishing utility for in-hospital mortality (AUC=0.604, *P*=0.087) and some merit for 30-day mortality (AUC=0.642, *P*=0.015). CURB-65 showed modest predictive value for in-hospital (AUC=0.715, *P*<0.0001) and 90-day mortality (AUC=0.672, *P*<0.0001), with the optimal derived cut-off CURB-65≥2.5 in our previous study^4^ had high specificity (75, 88% respectively) but low sensitivity (57, 46% respectively) (Table 2). The optimal derived cut-off for CURB-65 in this cohort was 3.5. The NEWS-2 showed higher predictive value for in-hospital (AUC=0.803, *P*<0.0001) and 90-day mortality (AUC=0.751, *P*<0.0001). The optimal cut-off for NEWS-2≥6.5 was found to have high sensitivity (73%) and specificity (72%) for predicting 90-day mortality, in contrast to high sensitivity (83%) and moderate specificity (63%) for predicting in-hospital mortality (Table 2).

**Table 2:** Outcomes of ROC curve analysis for evaluating optimal cut-off values for CURB-65, NEWS-2 and GAP scores, correlating with ARD-ILD mortality. Evaluation of the receiver operator curve (ROC) curve analysis for CURB-65, NEWS-2 and GAP scores correlate with 30- and 90-day mortality, in addition to in-hospital mortality.

|  | AUC | 95% CI | P-value | Decision Point | Sensitivity | Specificity | Youden's Index (J) |
| --- | --- | --- | --- | --- | --- | --- | --- |
| **SINGLE DISEASE SEVERITY SCORES** |  |  |  |  |  |  |  |
| **CURB-65** |  |  |  |  |  |  |  |
| In-hospital mortality | 0.715 | 0.620-0.810 | <0.0001 | 3.5 | 0.200 | 0.965 | 0.17 |
|  |  |  |  | 2.5 | 0.571 | 0.746 | 0.32 |
| 30-day mortality | 0.650 | 0.553-0.746 | 0.0021 | 1.5 | 0.476 | 0.737 | 0.21 |
| 90-day mortality | 0.672 | 0.591-0.753 | <0.0001 | 3.5 | 0.130 | 0.973 | 0.10 |
|  |  |  |  | 2.5 | 0.464 | 0.882 | 0.25 |
| **NEWS-2** |  |  |  |  |  |  |  |
| In-hospital mortality | 0.803 | 0.731-0.875 | <0.0001 | 6.5 | 0.829 | 0.634 | 0.46 |
| 30-day mortality | 0.750 | 0.669-0.831 | <0.0001 | 6.5 | 0.762 | 0.642 | 0.40 |
| 90-day mortality | 0.751 | 0.680-0.822 | <0.0001 | 6.5 | 0.725 | 0.718 | 0.44 |
| **GAP** |  |  |  |  |  |  |  |
| In-hospital mortality | 0.604 | 0.491-0.717 | 0.0869 | 4.5 | 0.607 | 0.554 | 0.16 |
| 30-day mortality | 0.642 | 0.541-0.744 | 0.0151 | 3.5 | 0.871 | 0.353 | 0.22 |
| 90-day mortality | 0.564 | 0.471-0.658 | 0.1887 | 4.5 | 0.545 | 0.568 | 0.11 |
| **COMBINATION DISEASE SEVERITY SCORES** |  |  |  |  |  |  |  |
| **CURB-65 and NEWS-2** |  |  |  |  |  |  |  |
| In-hospital mortality | 0.805 | 0.731-0.879 | <0.0001 | 0.157 | 0.857 | 0.620 | 0.48 |
| 30-day mortality | 0.681 | 0.589-0.822 | <0.0001 | 0.225 | 0.624 | 0.681 | 0.40 |
| 90-day mortality | 0.757 | 0.687-0.827 | <0.0001 | 0.392 | 0.710 | 0.682 | 0.39 |
| **CURB-65 and GAP** |  |  |  |  |  |  |  |
| In-hospital mortality | 0.764 | 0.671-0.856 | <0.0001 | 0.207 | 0.679 | 0.739 | 0.41 |
| 30-day mortality | 0.761 | 0.677-0.846 | <0.0001 | 0.140 | 0.903 | 0.504 | 0.41 |
| 90-day mortality | 0.727 | 0.646-0.809 | <0.0001 | 0.335 | 0.673 | 0.663 | 0.34 |
| **NEWS-2 and GAP** |  |  |  |  |  |  |  |
| In-hospital mortality | 0.802 | 0.745-0.895 | <0.0001 | 0.239 | 0.679 | 0.802 | 0.48 |
| 30-day mortality | 0.810 | 0.732-0.888 | <0.0001 | 0.177 | 0.839 | 0.639 | 0.48 |
| 90-day mortality | 0.752 | 0.675-0.828 | <0.0001 | 0.342 | 0.782 | 0.642 | 0.43 |

As the CURB-65 score correlated moderately to NEWS-2 score (R=0.552, *P*<0.0001), the combined ability of these two scores to predict mortality was assessed (Table 2). NEWS-2 retained significance to predict 90-day and in-hospital mortality after allowing for the effect of CURB-65 score (OR 1.34, *P*<0.0001, and OR 1.47, *P*<0.0001, respectively); however, CURB-65 was no longer a significant predictor after controlling for NEWS-2 for 90-day (OR 1.34, *P*=0.1648) or in-hospital mortality (OR 1.423, *P*=0.1791) (Supplementary Data 5). The addition of the GAP score improved the ability of the CURB-65 score to predict mortality (Table 2), with both GAP and CURB-65 retaining significance in each other’s presence. In contrast, the GAP score only improved the ability of the NEWS-2 score to predict 30-day mortality.

AUC, area under curve; CI, confidence interval; GAP, Gender, Age, Physiology Score; ROC, receiver operator characteristics. ROCs are provided within Supplementary Data 5 and 6.

### Secondary Outcomes

The median length of stay for patients admitted with an ARD-ILD was 6 days, but variability was observed across the cohort (IQR 3-11 days). Small numbers of patients received advanced respiratory support during their admission (n=29, 16%) although supplementary oxygen was frequently prescribed (n=159, 89%) (Table 1). ARD-ILD associated mortality was high; 20% of hospitalisations resulted in death, with 30- and 90-day mortality following admission of 24% and 38% (n=68) respectively (Table 1). Survival curve analysis indicated that patients admitted with ARD-IPF had higher in-hospital and 90-day mortality than patients with non-IPF-ARD (Figure 2). Univariate Cox regression analysis indicated that increasing NEWS-2 score was associated with higher mortality risk in-hospital and at 90- days; however, increased admission CURB-65 scores were not associated with an increased mortality risk (Table 3). Neither were higher admission CURB-65 nor NEWS-2 scores found to be associated with increased hazard of in-hospital mortality.

**Figure 2:**
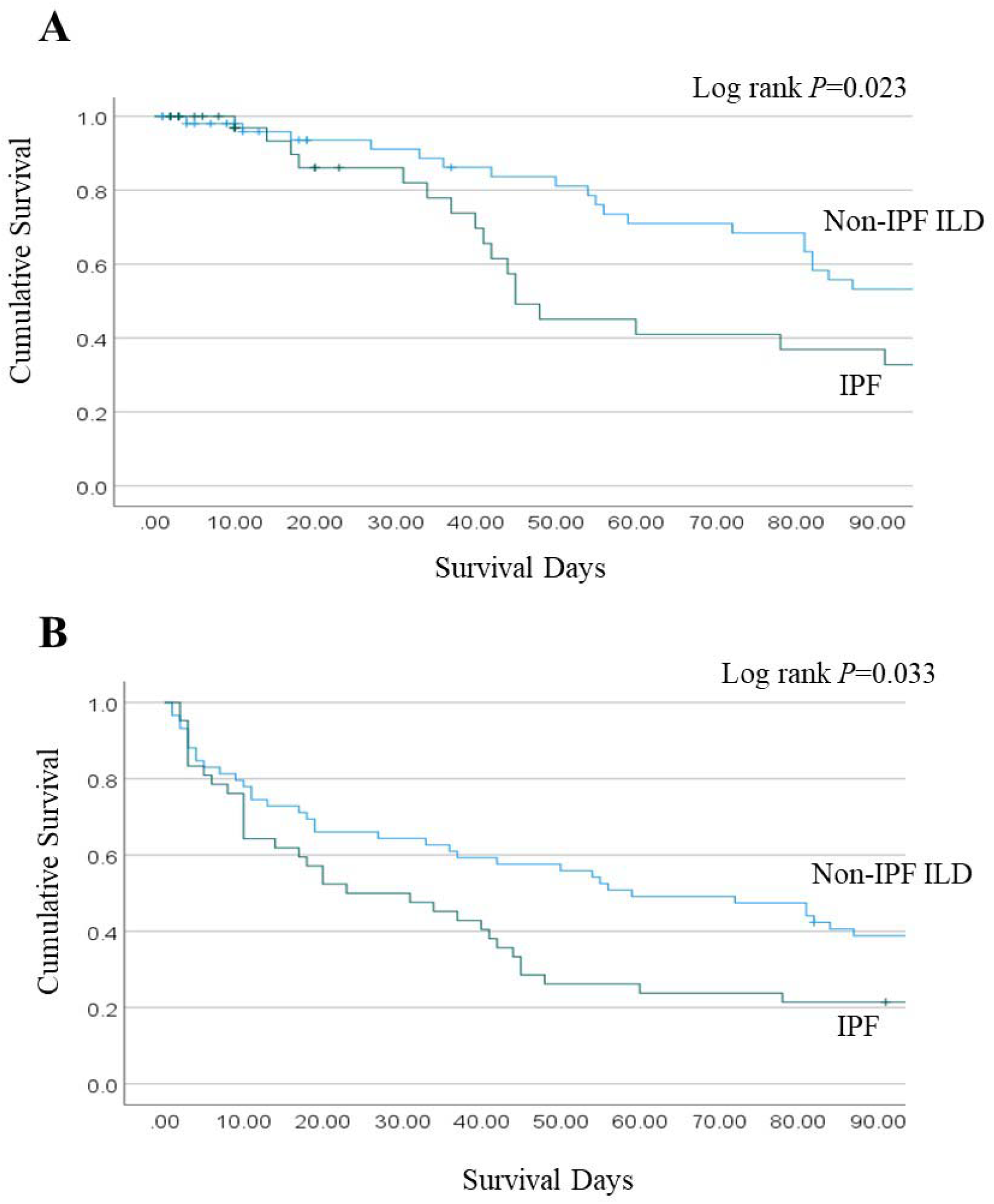
ARD-ILD survival curve analysis. Kaplan Meier survival curves for (A) in-hospital and (B) 90-day mortality, for patients admitted with ARD-IPF (green line) and non-IPF ARD-ILD (blue line).

**Table 3:** Association of baseline patient factors with in-hospital mortality and 90-day mortality following hospitalisation with ARD-ILD. Cox proportional hazard regression univariate analysis

| Analysis | Factor | Wald | HR | P-value | 95% CI |
| --- | --- | --- | --- | --- | --- |
| **In hospital mortality** | GAP score | 0.365 | 1.089 | 0.5452 | 0.826-1.435 |
|  | Smoking Status | 3.102 | 2.022 | 0.0778 | 0.924-4.428 |
|  | CURB-65 score | 3.163 | 1.565 | 0.0747 | 0.955-2.562 |
|  | NEWS-2 score | 3.841 | 1.188 | 0.0501 | 1.000-1.411 |
| **90-day mortality** | GAP score | 1.020 | 0.773 | 0.3132 | 0.469-1.274 |
|  | Smoking Status | 2.016 | 1.676 | 0.1558 | 0.822-3.419 |
|  | CURB-65 score | 1.089 | 0.753 | 0.2967 | 0.442-1.283 |
|  | NEWS-2 score | 4.251 | 1.231 | **0.0391** | 1.010-1.501 |

CI, confidence interval; GAP, Gender, Age, Physiology Score; HR, hazard ratio.

## DISCUSSION

The identification of patients with poor prognosis among those with ARD-ILD remains a significant challenge and there are limited data addressing the utility of illness severity scores to predict outcomes of these patients. This pragmatic, prospective study suggests that, as a predictor of mortality in this patient cohort, the admission NEWS-2 score has good discriminatory value for predicting in-hospital mortality and moderate discriminatory value for predicting 90-day mortality in patients following an ARD-ILD episode. As NEWS-2 is non-invasive and routinely calculated for all hospitalisations, this analysis suggests this severity score may be useful to non-specialists, enabling them to provide earlier appropriate care to ILD patients. To our knowledge, this is the first prospective evidence for utilising admission NEWS-2 as a tool to predict in-hospital and post hospitalisation morbidity following ARD-ILD. A previous retrospective cohort study conducted at one of the study hospitals found that both admission NEWS-2 and CURB-65 risk stratification tools were independent predictors of mortality in patients with IPF.^4^ The current study therefore confirms the findings of this previous retrospective cohort^4^ and hence admission NEWS-2 may represent a simple tool to help prognostication. Notably, the optimal derived cut-off value for admission NEWS-2 in predicting mortality across inpatient, 30- and 90-days was NEWS-2 ≥6.5, the same in both this prospective patient cohort and our previously reported retrospective cohort.^4^

The CURB-65 score at the point of admission was found to have little additional benefit for predicting mortality, either as an individual predictor or when used in exploratory analyses with NEWS-2. Furthermore, we found that admission CURB-65 was able to predict both 90- day and in-hospital mortality in ARD-ILD with high specificity but low sensitivity, in keeping with previous data from our IPF cohort^4^. Aligning with these findings, Yamazaki *et al* found that CURB-65, Pneumonia Severity Index (PSI) and Sequential Organ Failure Assessment (qSOFA) were all predictive of inpatient mortality from pneumonia in a retrospective study of 79 patients with IPF.^21^ In that study, the optimal CURB-65 cut-off value was ≥3.0, differing slightly from the value derived in our previous retrospective IPF cohort,^4^ but contrasting with the optimal cut-off ≥2.5 derived in this current prospective ILD cohort. Notably, the admission CURB-65 score did not add value to the ability of the NEWS-2 score to predict poor outcomes. In contrast, exploratory analyses indicate there is some evidence of increased predictive ability on outcomes when NEWS-2 and GAP scores are used in tandem. Overall, these findings from different centres and patient populations suggest that the disease severity scores show promise in predicting poor outcome following hospitalisation with acute respiratory deterioration.

As reported before the emergence of SARS-CoV-2, patients hospitalised with ARD-IPF had a poor prognosis.^4,22–24^ We found that ARD-ILD continues to cause a significant disease burden in secondary care following SARS-CoV-2 emergence, with a median hospital admission of 6 days (IQR3-11). Whilst we found in-hospital and 90-day mortality rates were significantly higher in the IPF group than the non-IPF group, all admissions related to an ARD were associated with significant mortality; 24% and 38% for 30- and 90-day mortality, respectively. The median survival in this cohort was 23 days, slightly shorter than the survival times in the literature ranging between 1 and 4 months after AE-IPF.^25–28^ In keeping with our findings, Suzuki *et al* reported that AE IPF was associated with a 47% 90-day mortality, which was statistically poorer than non-IPF AE 90-day mortality of 38%.^29^ Other studies have reported IPF patients have a significantly shorter overall and post-hospitalization survival time following an acute respiratory deterioration compared with other ILDs.^22,29^ Our in-hospital mortality data supports early and frank discussions between patients/families and clinicians surrounding the high mortality associated with ARD, even in the context of potentially ‘treatable’ causes e.g. infection and pulmonary embolism. Furthermore, the high short-term mortality after discharge may prompt clinicians to consider early discussions surrounding palliation and/or transplantation in suitable candidates. In practice, the treatment decisions in patients with ARD-ILD faced by both specialist and non-specialist clinicians, patients and their families are profound and often irreversible. Making these decisions optimally requires the integration of many differing and individual considerations, including accurate prognostic information, and as the predictive characteristics of the scores as determined in this study may not add value above the ability of experienced clinicians to advise patients and their families. Whilst antifibrotics reduce mortality in ILD, to our knowledge it is currently unknown what the impact of antifibrotics on severity of AE ILD once it occurs;^30^ although, nintedanib may reduce severity of first exacerbation.^31^ The NICE (National Institute for Health and Care Excellence) technology appraisal guideline for antifibrotics in non-IPF ILD did not exist at the time this cohort was hospitalised. We note that 57% of IPF patients in this study were receiving antifibrotic treatment at the time of admission, but this analysis would not be powered to determine if antifibrotic use in IPF reduces the severity of AE-IPF in patients hospitalised with this condition.

There were few admissions secondary to AE-ILD, which prevented further detailed analyses of any differences in the mortality between AE-ILD and other parenchymal causes of ARD. Teramachi *et al* showed 90-day mortality of AE-IPF patients was significantly higher than ARD due to other parenchymal causes (46% [16/35] vs 17% [12/71] respectively; *P*=0.002).^14^ It is not possible to ascertain fully to what extent healthcare access and patient behaviour during the pandemic affected these outcomes. The Task Force for Lung Health suggested that over a third of people with pre-existing lung problems felt pressure to avoid or delay seeking treatment.^32^ Hence, patients may have delayed presentation to hospital. Alternatively, the emergence of a novel pathogen may have affected outcomes of patients with ARD, either directly or indirectly through changes in the epidemiology of other acute respiratory infection, treatment pathways or other aspects of patient healthcare provision. In non-ILD patients hospitalised with SARS-CoV-2 infection, admission NEWS-2 correlates moderately well with severe outcomes such as ITU admission, positive airway pressure support or death; however, there were no such significant correlations for admission CURB-65 (*P*>0.05).^33^

This study has many strengths. It was undertaken as a prospective two site cohort study which screened hospital admissions for signs/symptoms of acute respiratory disease. This study therefore does not rely on ICD-10 coding or solely on data-linkage. There were minimal missing data, and the study includes adults who lack capacity to consent through a consultee and by specific authorisation to use certain data without consent, thereby ensuring full ascertainment of ARD-ILD during this period. The medical records were linked with community records to obtain detailed and accurate data for each study participant.

There are also some limitations of this study. We assessed the use of admission severity scores as mortality predictors in patients hospitalised with ARD-ILD and this analysis cannot and does not assess their usefulness in ILD patients in stable state. We assessed ARD-ILD at both acute care NHS hospitals in Bristol and 83% of this cohort is Caucasian; therefore, we cannot be sure that results are generalisable to other patient populations. We are not sufficiently power to statistically test whether two correlated area under the curves differ when one element is in common (CURB-65 and GAP vs. NEWS and GAP) or whether one model nested in another (CURB-65 versus CURB-65 and GAP) differ in terms of their AUROC. This study used linked national healthcare records to determine mortality, and did not ascertain the specific cause of death. We acknowledge that the ILD exacerbation rate in this cohort appears to be low, we were therefore unable to confirm our previous findings that mortality associated with ARD-ILD was high, irrespective of the underlying cause for the deterioration. Notably only three patients met the definition of AE-ILD. Not all patients with evidence of pneumonia had a CT performed and it is therefore possible that some of these patients had a triggered exacerbation with increased ground glass opacification not evident on CXR. We were also unable to determine the time from ILD diagnosis until hospitalisation with ARD, and therefore could not adjust for this co-variate. The impact of shielding during the COVID pandemic on exacerbation rate is a further factor to be considered. Lastly, it is difficult to determine whether healthcare access may have affected time to hospitalisation and hence outcomes, as previously discussed. Others have reported that patients with chronic respiratory disease had fewer admissions to secondary-care during the COVID-19 pandemic^32,34^ and this may have affected the results of this study, although the direction of effect is unclear.

## CONCLUSION

Simple illness severity scores may permit refinement of ARD management of ILD patients and if survival to discharge is achieved, permit early discussion with patients, referral to transplantation or palliative care planning as appropriate.

## Data Availability

The data used in this study are sensitive and cannot be made publicly available without breaching patient confidentiality regulations. Therefore, individual participant data and a data dictionary are not available to other researchers.

## ABBERVIATIONS

6MWT: c6-minute walk test
aLRTD: acute lower respiratory tract disease
AE: acute exacerbation
ARD: Acute respiratory deterioration
AUROC: Area Under the Receiver Operating Curve
CRP: C-reactive protein
CI: confidence interval
CCI: Charlson comorbidity index
CPFE: combined pulmonary fibrosis and emphysema (IPF with emphysema)
CT: computerized tomography (CT) scan
CTD: connective tissue disease
CXR: Chest x-ray (i.e. chest radiograph)
GAP: Gender-Age-Physiology
FVC: Forced vital capacity
HFNO: high flow nasal oxygen
HR: hazard ratio
ILD: Interstitial lung disease
IMV: invasive mechanical ventilation
IPF: Idiopathic pulmonary fibrosis
IQR: interquartile range;
LFTs: lung function tests
LOS: length of stay
MDT: multidisciplinary team
NEWS-2: National Early Warning Score-2
NIV: non-invasive ventilation
NSIP: non-specific interstitial pneumonia
NP-LRTI: non-pneumonic lower respiratory tract infection
OR: odds ratio
PFTs: pulmonary function tests
PSI: pneumonia severity index
qSOFA: Sequential Organ Failure Assessment
ROC: Receiver Operating Curve
TLCO: transfer capacity of the lung, for the uptake of carbon monoxide

## AUTHOR CONTRIBUTIONS

CH, RW, PW, AF and SLB generated the research question and analysis plan. The data for this study was collected by the AvonCAP Research Team, CH, RW, JR, MGG and ZSB. CH, MGG, ZSB and RW verified the data. RW, CH, PW, AF and SLB undertook data analysis.

All authors were involved in the final manuscript preparation and its revisions before publication. AF and SB provided oversight of the research.

## ACKNOWLEDGEMENTS

The authors would like to acknowledge the Research Teams at North Bristol NHS and University Hospitals Bristol and Weston NHS Trusts, including Helen Lewis-White, Diana Benton, Anna Morley, Jade King and Nicola Manning. Furthermore, we would like to acknowledge the significant contribution to the AvonCAP study made by Rachel Davies, Adam Taylor, Mai Baquedano and the IT team, including Stuart Robinson, at North Bristol NHS Trust.

## The AvonCAP Research Group

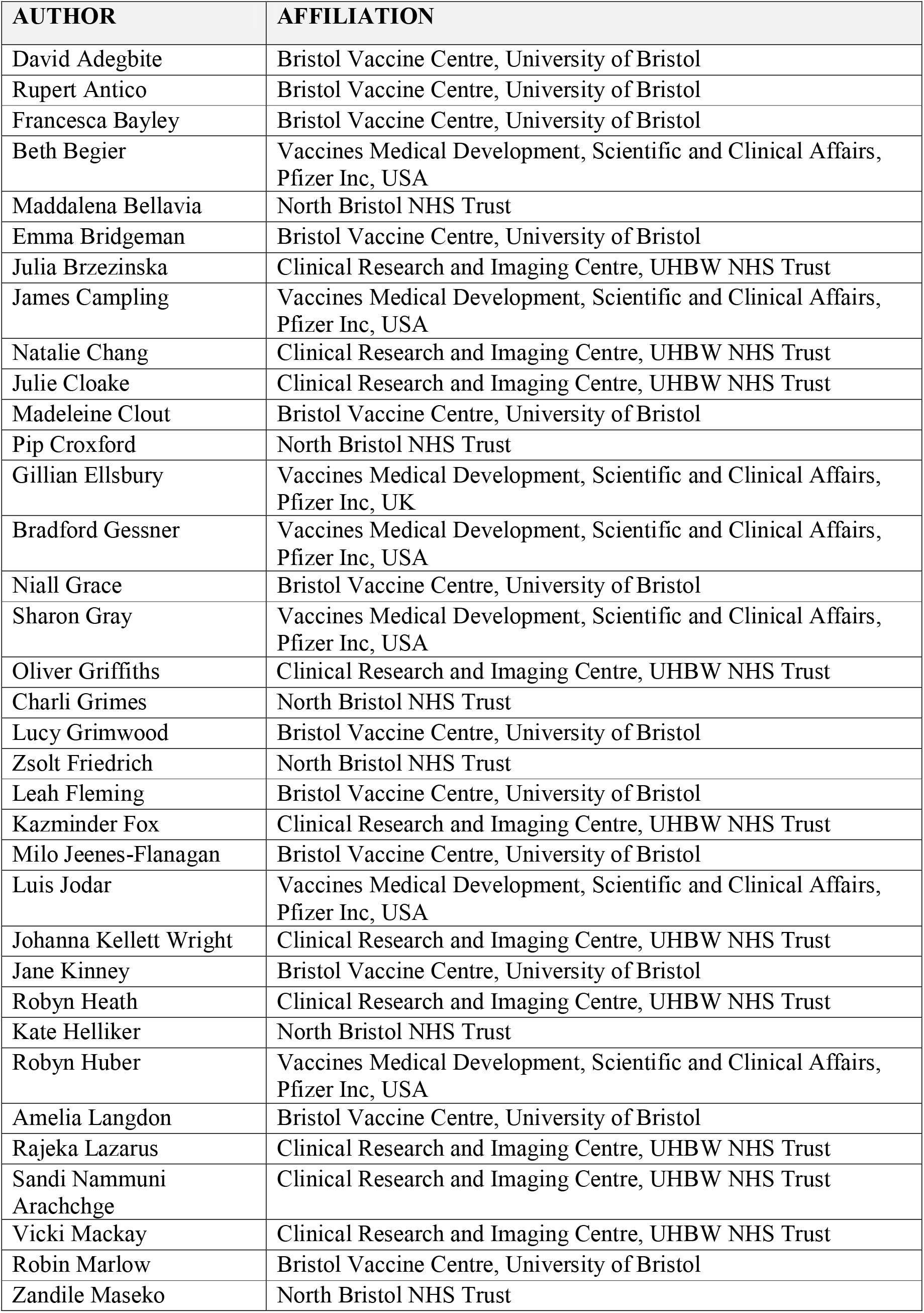

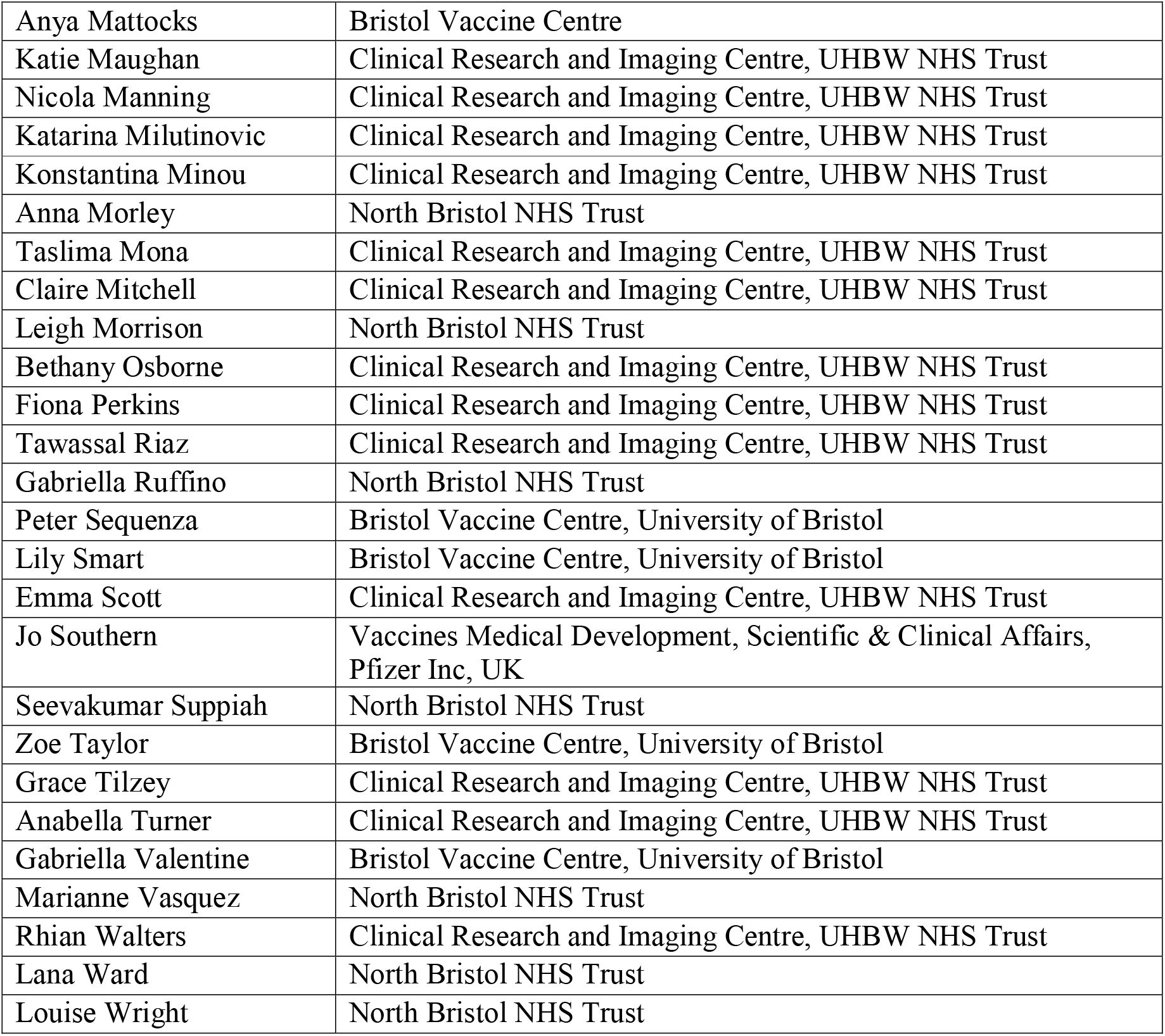

**Supplemental Data 1:**
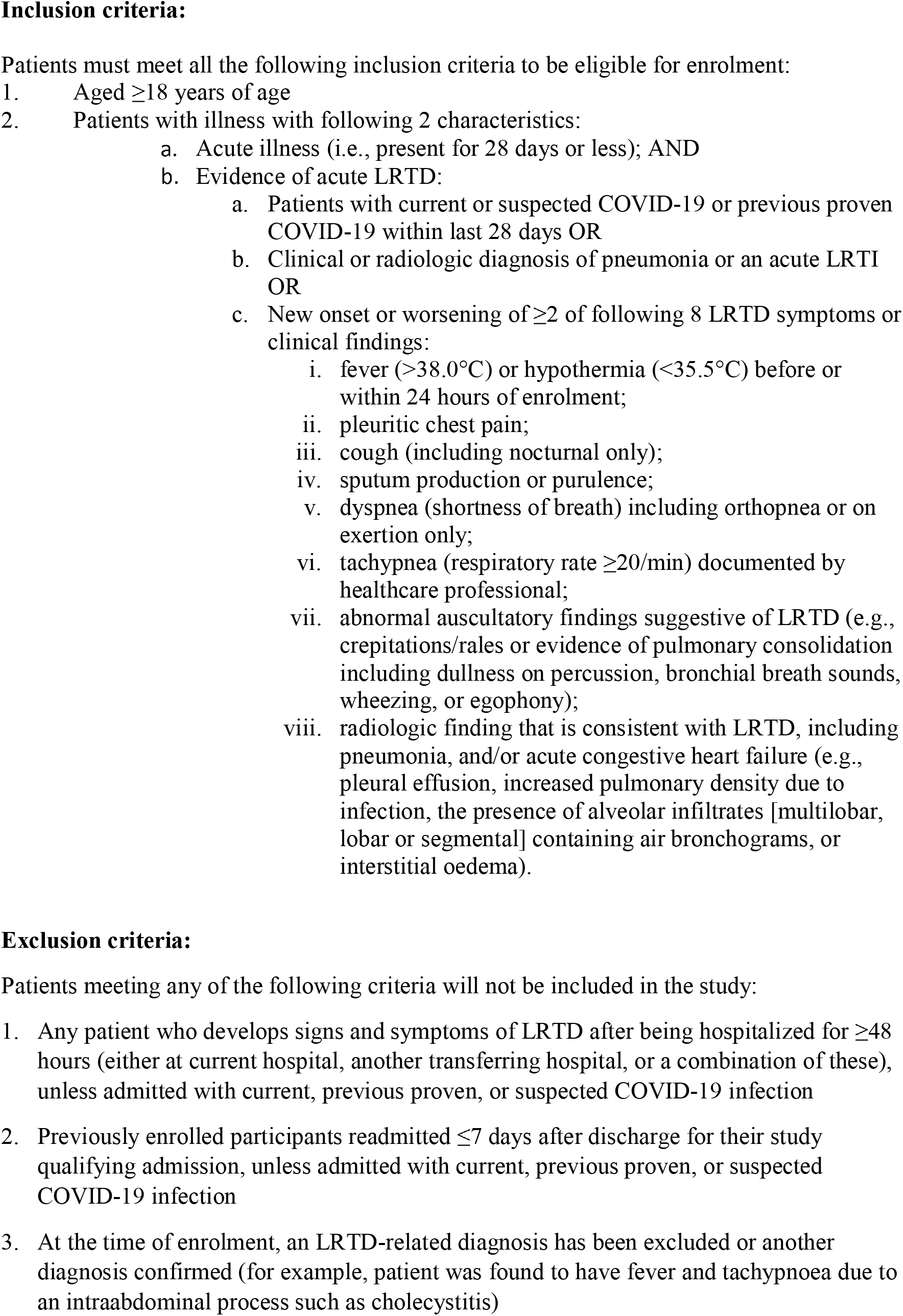
AvonCAP inclusion and exclusion criteria.

**Supplemental Data 2:**
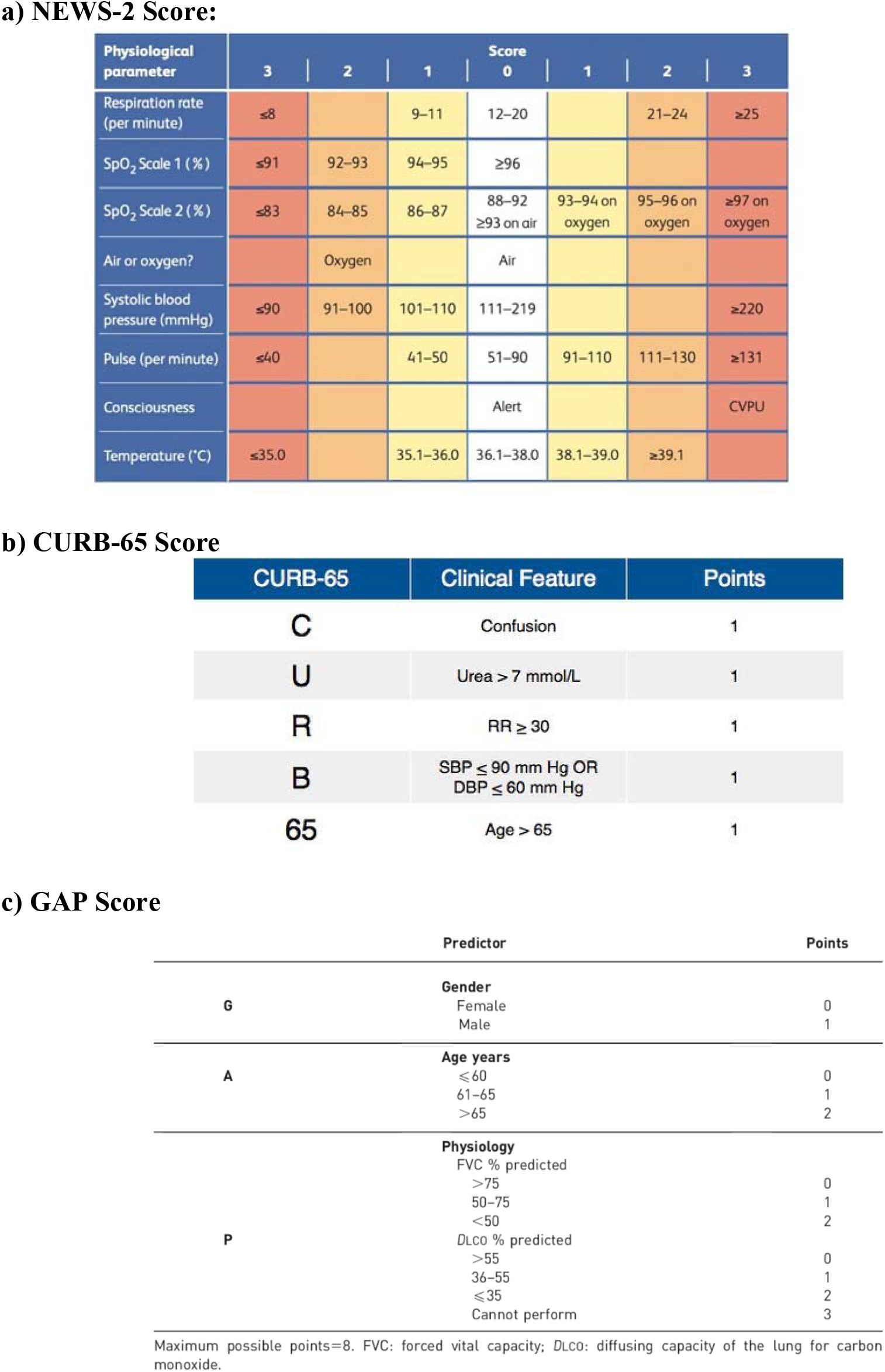
Disease severity scores.

**Supplementary Data 3:**
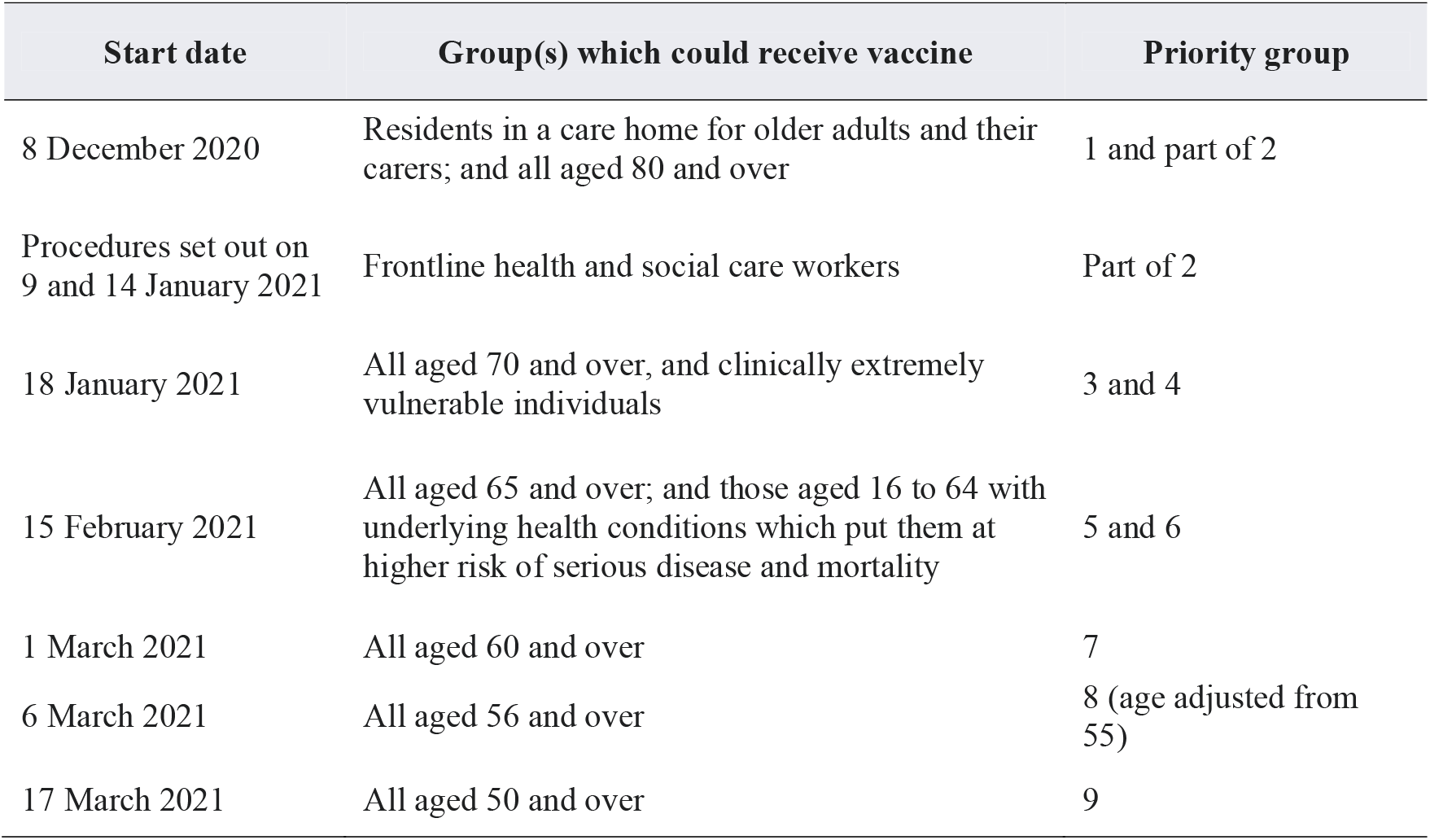
Availability of vaccine appointments in England by date.

**Supplementary Data 4:**
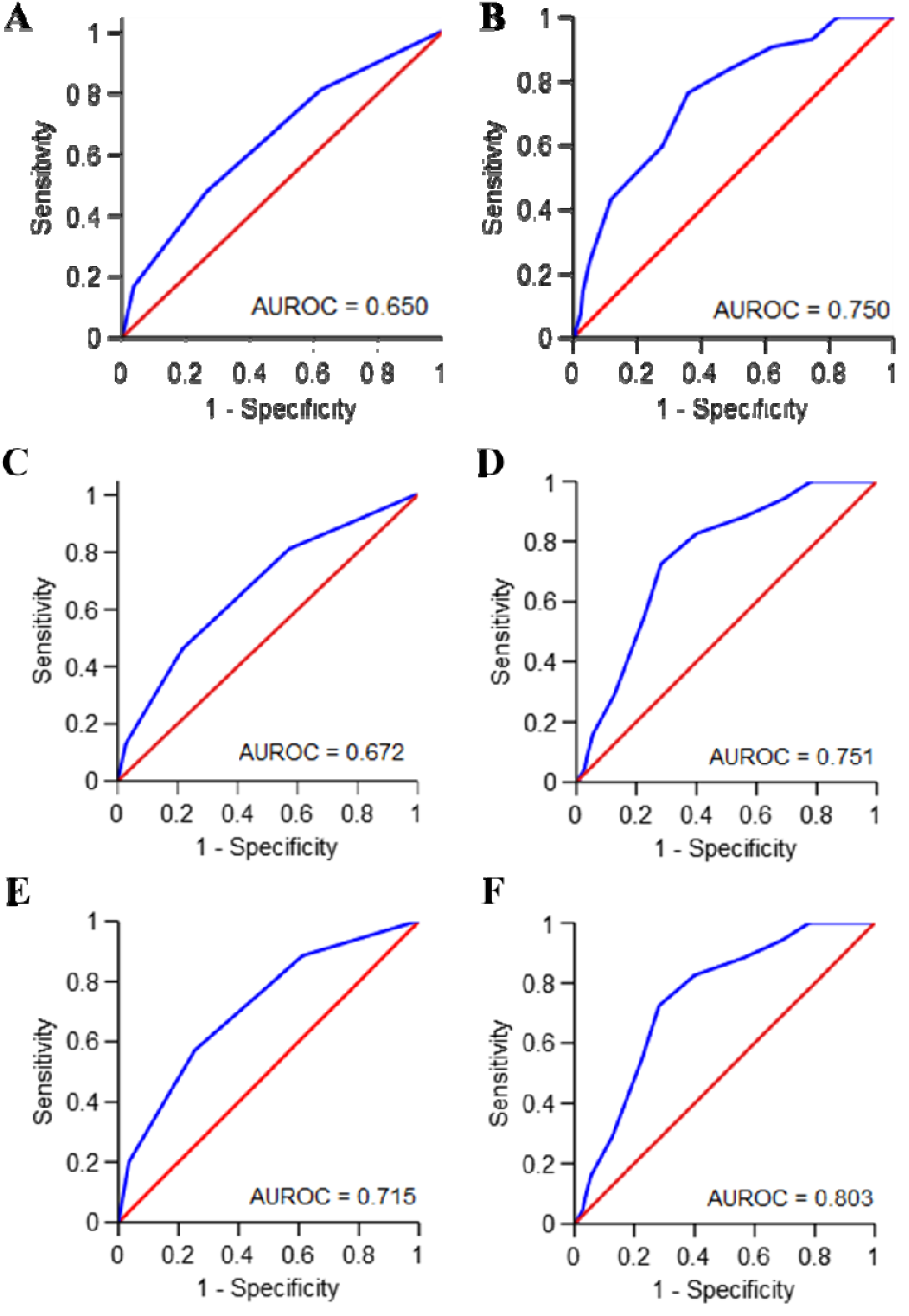
Receiver Operator Characteristic (ROC) curves. ROC curves for 30-day mortality and (A) CURB-65 score and (B) NEWS-2 score; 90-day mortality and (C) CURB-65 and (D) NEWS-2 score; and, in-hospital mortality and (E) CURB-65 and (F) NEWS-2.

**Supplementary Data 5:**
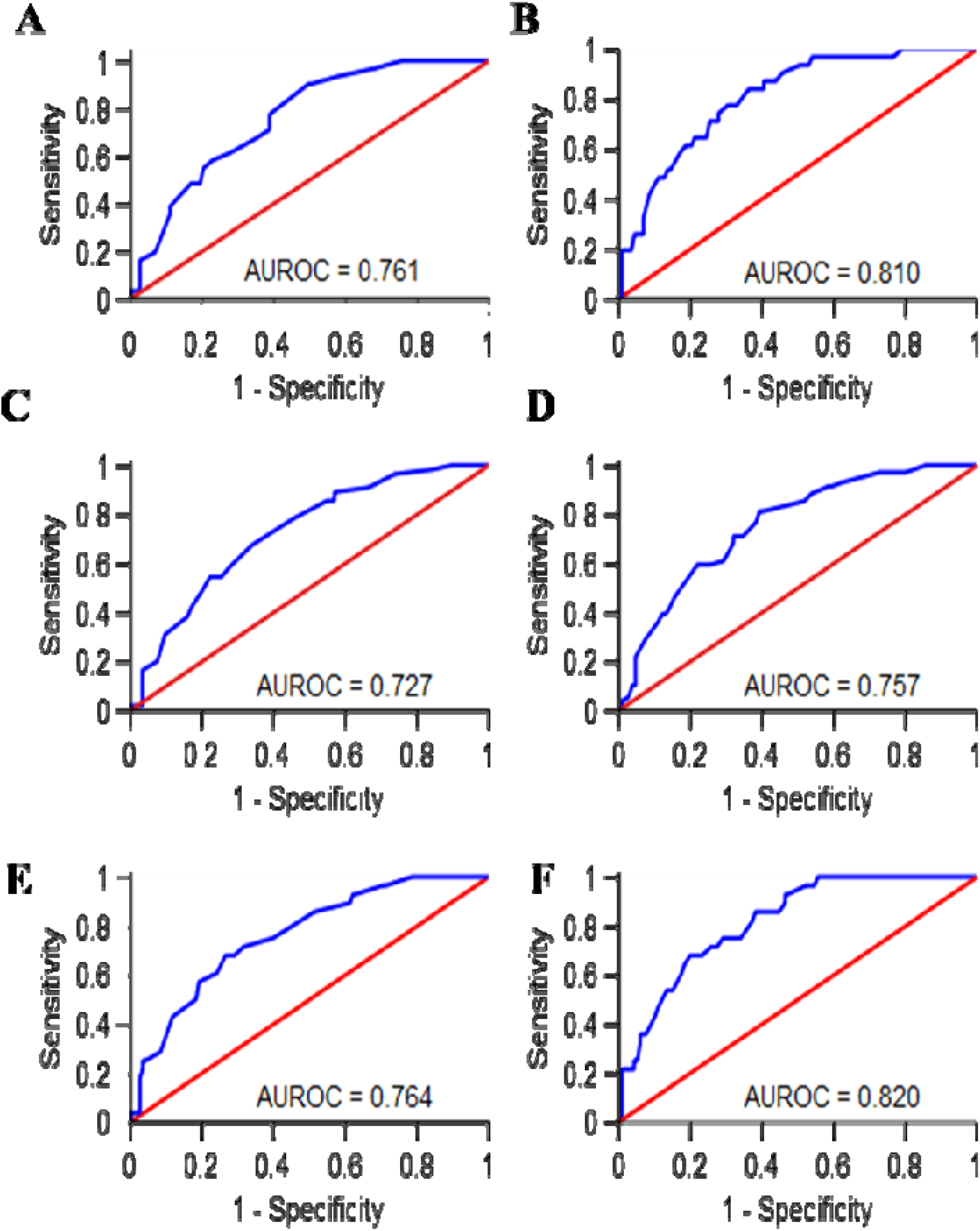
Receiver Operator Curves (ROC) for NEWS-2 and CURB-65 combined with GAP score. ROC curves for 30-day mortality and GAP with (A) CURB-65 score and (B) NEWS-2 score; 90-day mortality and GAP with (C) CURB-65 and (D) NEWS-2 score; and, in-hospital mortality and GAP with (E) CURB-65 and (F) NEWS-2.

**Supplementary Data 6:**
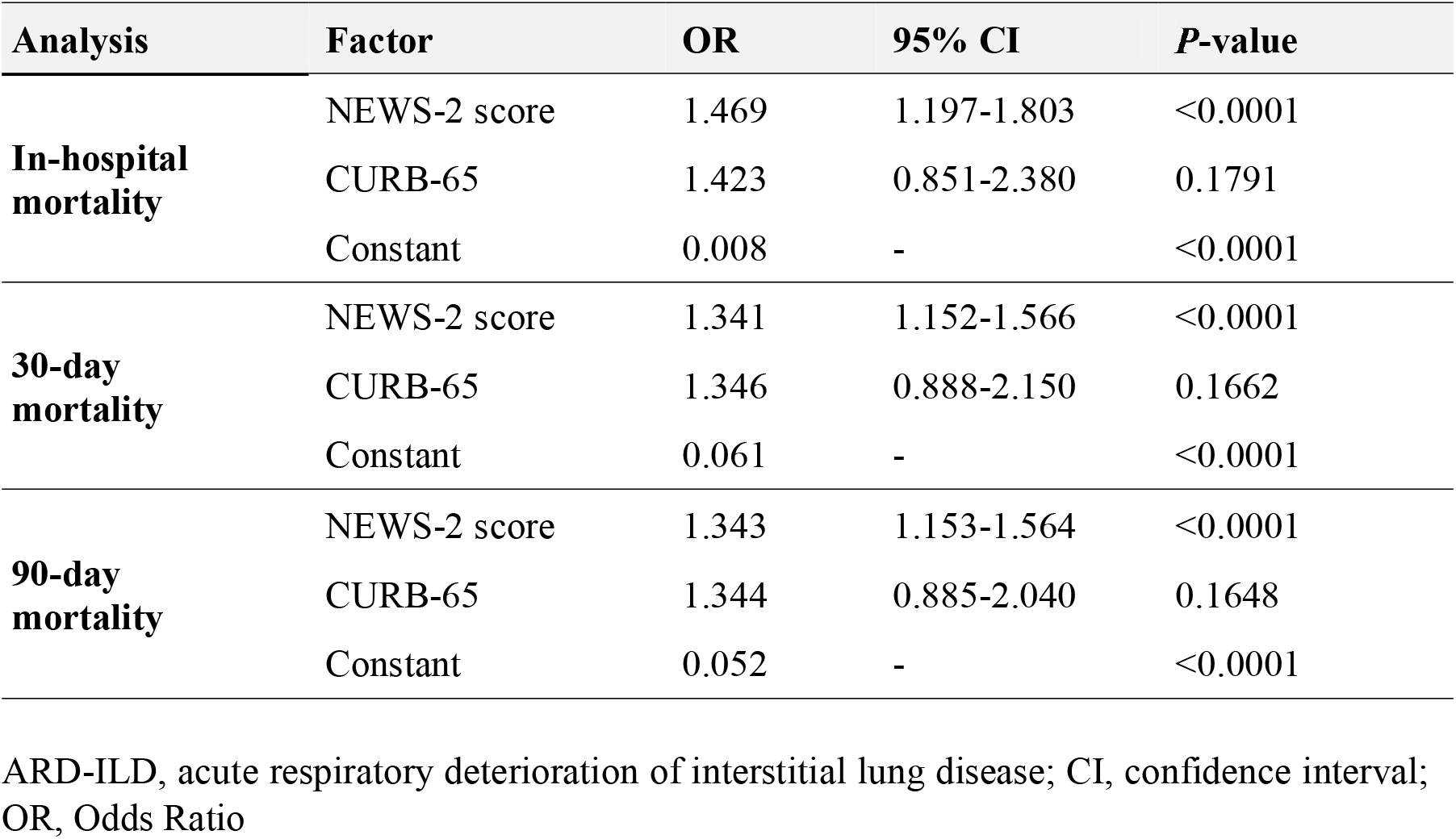
Joint predictive value of NEWS-2 and CURB-65 scores in predicting in-hospital mortality and 90-day mortality following hospitalisation with ARD-ILD. (A) Binary logistic regression analysis for joint predictive value of NEWS-2 and CURB-65 scores in predicting mortality following hospital admission with ARD-ILD.

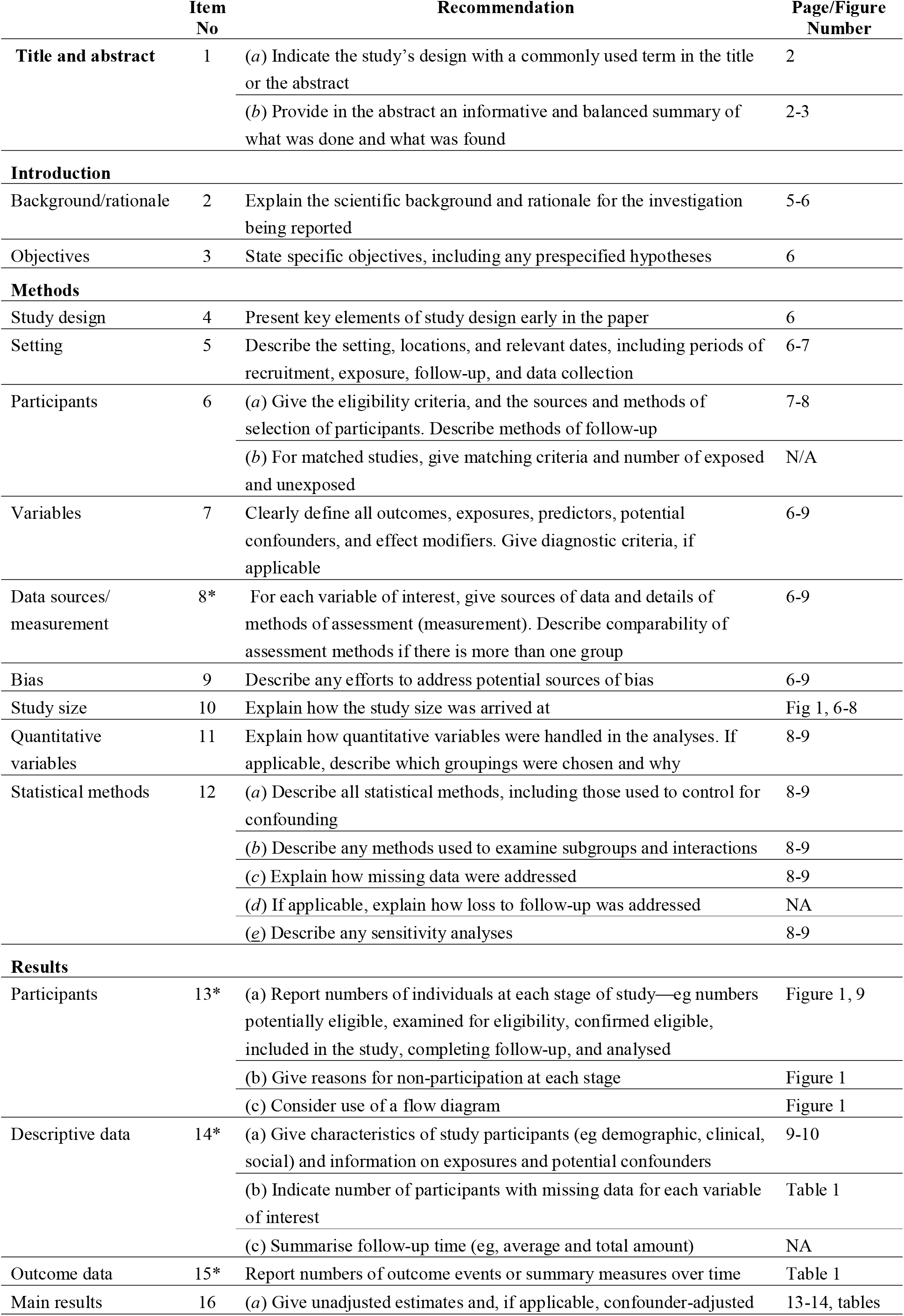

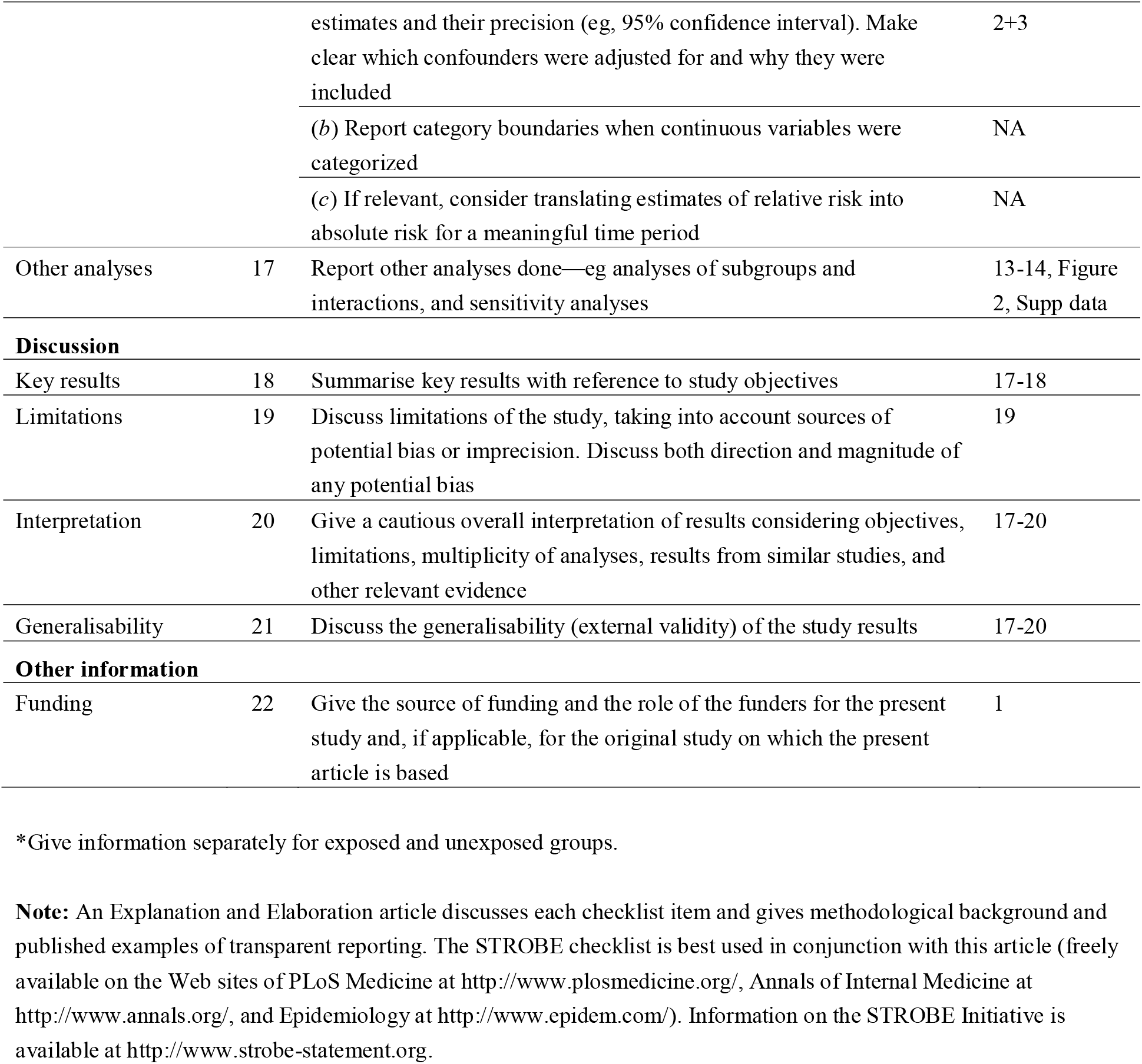
STROBE Statement—Checklist of items that should be included in reports of ***cohort studies***

